# Screen Time as a factor for Attention Deficit Hyperactivity Disorder (ADHD) in children: A Systematic Review

**DOI:** 10.1101/2025.04.29.25325745

**Authors:** Um ul Baneen Zehra, Roshanay Raza, Emaan Tindyala, Karthik Kanna Venkatesh, Iram Mansoor, Mehar Zehra, Ali Ejaz, Fateema Tanveer, Muhammad Bin Aslam, Sadia Rekhum

## Abstract

This study examined the relationship between children’s screen time and the potential development of Attention Deficit Hyperactivity Disorder (ADHD). Through analysis of various credible sources over the past decade, including Google Scholar and PubMed Central, a significant correlation between increased screen time and the exacerbation of ADHD symptoms was identified. The research suggested that prolonged screen exposure heightened the risk of developing ADHD and intensified its severity. Additionally, excessive screen usage was found to disrupt sleep patterns and undermine healthy eating habits. This review emphasized the importance of effectively managing ADHD symptoms to potentially mitigate the likelihood of its onset and enhance the overall well-being of children diagnosed with the disorder.

## Introduction and Background

In contemporary society, smartphones, televisions, and tablets have become integral to the daily lives of both children and young adults. These devices serve various purposes, facilitating both learning and entertainment. Technological advancements continuously enhance our engagement with screens, offering a plethora of activities available almost at any time and location. However, it is not advisable to exceed a screen time of two hours per day (Lissak, 2018). Research indicates that adolescents possessing smartphones tend to engage with screens more frequently than their peers with conventional mobile phones (Brem et al., 2014). A study conducted by the American Academy of Pediatrics revealed a concerning trend regarding the increased use of digital devices among children across numerous countries, including those in the West and the Far East. Specifically, in 2011, 52% of children aged 0 to 8 had access to mobile devices, whereas, by 2013, this figure escalated to 75% (Chassiakos et al., 2016).

Attention Deficit Hyperactivity Disorder (ADHD) is a prevalent concern among children and adolescents, with a prevalence rate of approximately 5% within this population (Drechsler et al., 2020). This condition presents significant challenges for affected individuals and their families, often complicating aspects such as mental health, self-esteem, and emotional regulation, thereby hindering daily activities (Klassen et al., 2004). Recent studies suggest that ADHD may persist into adulthood, resulting in ongoing difficulties with motor function and other related issues (Zalsman and Shilton, 2016). Further research is warranted to elucidate the factors contributing to the onset of ADHD in adults.

While digital devices undoubtedly offer notable advantages, they are associated with various health and mental health concerns, including ADHD. A study in 2018 indicated that prolonged screen time could be linked to sleep disturbances and depressive symptoms, which may, in turn, exacerbate the prevalence of disorders such as ADHD (Lissak, 2018). Engaging in substantial screen time, particularly with exposure to violent or fast-paced content, may activate neural pathways associated with pleasure chemicals, potentially resulting in symptoms that are misattributed to ADHD. Reducing screen time has been shown to alleviate these symptoms.

## Methodology

This systematic review strictly adhered to the PRISMA (Preferred Reporting Items for Systematic Reviews and Meta-Analyses) protocol to ensure rigor and consistency in the research design. Notably, no public input was solicited during its development.

Attention Deficit Hyperactivity Disorder (ADHD) in children is recognized as a multifactorial neurodevelopmental condition. This review encompasses studies that investigate the relationship between ADHD in children and their recreational screen time habits.

The methodology focused exclusively on full-text journal articles, with the inclusion criteria restricted to publications in the English language. Studies were excluded if they did not examine the link between ADHD and screen time, were limited to abstract-only format, or involved non-human research subjects.

The primary outcomes of the study were centered on the increasing rates of ADHD diagnoses among children with excessive screen time exposure, as well as the correlation between the duration of screen time and the incidence of ADHD in childhood.

A thorough screening and data extraction process was conducted utilizing the PubMed Central and Google Scholar databases to identify pertinent studies published between 2013 and 2023 that pertain to ADHD. The search criteria and strategy are summarized in Table 1. In total, 2,509 articles were identified through this process. After a meticulous review, 22 articles met the established inclusion criteria for this systematic review, as illustrated in Figure 1.

**Figure 1:**
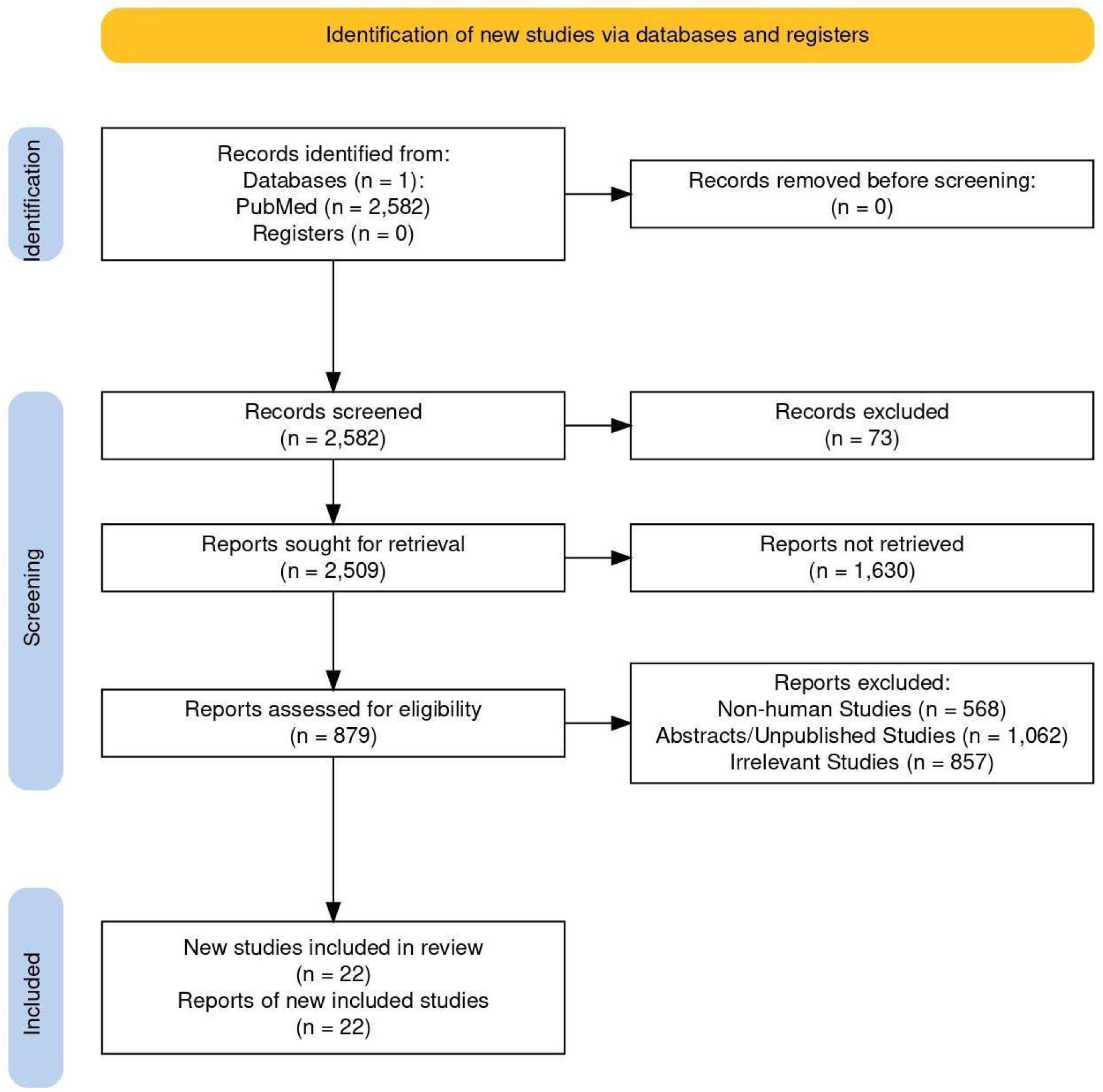
PRISMA flowchart for the selection of studies

**Table 1:** Search Criteria.

| **Databases** | **PubMed** |
| --- | --- |
| Keywords | ADHD, Screen time, Children, Social Media, Hyperactivity |
| Publications | Only English language article, human studies and full text articles were included. |
| Time frame | 2013-2023 |

The data extracted from the selected studies were systematically organized into a comprehensive table, which detailed each study according to four key parameters: database source, authorship, year of publication, and study findings specifically elucidating the relationship between ADHD and screen time exposure, as shown in Table 2.

**Table 2:** Review of articles.

| Database | Author | Year | Results |
| --- | --- | --- | --- |
| Pubmed | Sage Pickren | 2022 | Parents report an increase in children's screen time by 1 to 3 and worsening of ADHD symptoms<sup>[7]</sup> |
| PubMed | Monique<br>Moore Hill | 2020 | 36 month old children with most screen time had elevated ADHD symptoms<sup>[8]</sup> |
| PubMed | Birgit Levelink | 2021 | There was a weak relationship between hours spent watching television and ADHD symptoms<sup>[9]</sup> |
| PubMed | Udena<br>Ruwindu<br>Attygalle | 2020 | Study shows no association between ADHD and screen time<sup>[10]</sup> |
| PubMed | Anyi Yang | 2022 | Study shows that there are shared neuronal overlaps of ADHD and increase screen time and provide evidence that polygenic risk of ADHD is related to increased screen time<sup>[11]</sup> |
| PubMed | Katie N. Paulich | 2021 | Screen time is moderately associated with behavioral problems including ADHD, poor academic performance and poor sleep<sup>[12]</sup> |
| PubMed | Lian Tong | 2016 | Study shows that children with ADHD symptoms have more unhealthy behavior including increased screen time, sedentary lifestyle and poor eating habits<sup>[13]</sup> |
| PubMed | Ilaria Montagni | 2016 | Higher electronic screen exposure is associated with self-perceived attention and hyperactivity problems<sup>[14]</sup> |
| Pubmed | Gadi Lissak | 2018 | Study correlates increased screen time with neurological adverse health consequences including ADHD symptoms<sup>[1]</sup> |
| Pubmed | Pedro San<br>Martin Soares | 2021 | ADHD symptoms at 22 years were associated with television time at 11 years, computer time at 18 years and total screen time at ages 11, 15, and 18 years<sup>[15]</sup> |
| Pubmed | Sivapriya<br>Vaidyanathan | 2021 | Preschoolers diagnosed with ADHD had screen time more than recommended duration<sup>[16]</sup> |
| Pubmed | Tony Xing<br>Tang | 2022 | Increased screen time was a risk for inattention and hyperactivity or impulsivity behaviors in urban children<sup>[17]</sup> |
| Pubmed | Sukhpreet K.<br>Tamana | 2019 | Increased screen-time in pre-school is associated with worsening of inattention<sup>[18]</sup> |
| Pubmed | Vanessa<br>K.Thoma | 2020 | Increased media time and disturbed sleep-wake behavior increased the risk of ADHD-like symptoms<sup>[19]</sup> |
| Pubmed | Laura Masi | 2021 | Study shows correlation between the severity of ADHD symptoms and excessive use of video games<sup>[20]</sup> |
| Pubmed | Eddy Cavalli | 2021 | Study shows a correlation between evening TV exposure and higher SDSC and ADHD scores and exacerbation of ADHD symptoms by increased screen time indirectly due to sleep disturbances<sup>[21]</sup> |
| Pubmed | Lan Shuai | 2021 | ADHD children with problematic social media use have more severe symptoms<sup>[22]</sup> |
| Pubmed | Julia Hansen | 2022 | German children meeting all the guidelines of screen time, physical activity and sleep had less frequent ADHD and depressive symptoms<sup>[23]</sup> |
| Pubmed | Katherine H. Peppers | 2020 | Reduced screen use before sleep can improve sleep hygiene and decrease ADHD symptoms<sup>[24]</sup> |
| Pubmed | Franzika<br>Waller | 2023 | Study shows direct association between ADHD symptoms and screen time<sup>[25]</sup> |
| Pubmed | Lisa B. Thorell | 2022 | ADHD patients appear more vulnerable to developing high or problematic use of digital media, and digital media also has effects on severity of ADHD symptoms<sup>[26]</sup> |
| Pubmed | Matthew<br>M.Engelhard | 2019 | Study identifies that individuals with ADHD are sensitive to screen media technology and focuses on quantifying screen media technology<sup>[27]</sup> |

## Results

Several studies have investigated the relationship between screen time and Attention Deficit Hyperactivity Disorder (ADHD), yielding mixed findings. Pickren et al. (2022) reported that parents noted an increase in their children’s screen time by 1 to 3 hours, which correlated with a deterioration in ADHD symptoms. Similarly, Hill et al. (2020) found that 36-month-old children with the highest screen time exhibited elevated symptoms of ADHD. Conversely, Levelink et al. (2021) observed a weak correlation between television exposure and ADHD symptoms, suggesting a limited impact of screen time on the manifestation of ADHD. In contrast, Attygalle et al. (2020) found no significant association between ADHD and screen time, indicating that screen time may not be a central factor in the development of ADHD symptoms.

Yang et al. (2022) highlighted shared neuronal overlaps between ADHD and increased screen time, proposing that the polygenic risk associated with ADHD could be linked to higher screen time exposure. Paulich et al. (2021) established a moderate association between screen time and various behavioral issues, including ADHD, poor academic performance, and sleep disturbances. In a study conducted by Tong et al. (2016), children exhibiting ADHD symptoms were reported to engage in unhealthy behaviors, such as increased screen time, sedentary habits, and poor dietary practices. Montagni et al. further supported the connection between screen time and ADHD, noting that higher exposure to electronic screens was associated with self-reported attention and hyperactivity problems.

Lissak (2018) identified a correlation between increased screen time and adverse neurological health consequences, including symptoms of ADHD, suggesting that excessive screen exposure could contribute to the development of these symptoms. The observational study by Soares et al. (2021) found that ADHD symptoms at the age of 22 were linked to television and computer time during adolescence, emphasizing the long-term effects of screen time on ADHD. Furthermore, a cross-sectional exploratory study conducted in South India indicated that preschoolers diagnosed with ADHD were more likely to exceed the recommended duration of screen time, presenting a potential risk factor for exacerbating symptoms (Vaidyanathan et al. 2021). Tang et al. (2022) noted that increased screen time among urban children was a risk factor for inattention, hyperactivity, and impulsive behavior.

Observations from studies by Tamana et al. (2019) and Waller et al. (2023) demonstrated that increased screen time in children was associated with a worsening of inattention, supporting the notion that early exposure to screens may exacerbate ADHD symptoms. Thoma et al. (2020) concluded that an increase in media consumption and disrupted sleep-wake patterns elevated the risk of ADHD-like symptoms, suggesting the influence of sleep disturbances on screen time-related ADHD. The literature also reports a correlation between the severity of ADHD symptoms and excessive video game use, further underscoring the impact of digital media on ADHD (Masi et al.) (2021)).

Cavalli et al. (2021) established a connection between evening television exposure and elevated ADHD scores, asserting that sleep disturbances may exacerbate symptoms, thereby highlighting an indirect effect of screen time on ADHD. Shuai et al. (2021) demonstrated that children with ADHD who engaged in problematic social media use displayed more severe symptoms, indicating a significant relationship between social media habits and the intensity of ADHD symptoms. Hansen et al. (2022) reported that German children who adhered to established guidelines for screen time, physical activity, and sleep experienced fewer symptoms of ADHD and depression, suggesting that balanced lifestyle habits may mitigate the manifestation of ADHD symptoms.

Peppers et al. (2020) demonstrated improvements in both sleep hygiene and ADHD symptoms through the reduction of screen time in their study population, yielding results with external validity that further underscore the importance of managing screen time for the effective control of symptoms. Individuals with ADHD may possess a heightened susceptibility to developing excessive or problematic digital media use, which could exacerbate the severity of their symptoms, thus perpetuating a detrimental cycle (Thorell et al. 2022). Additionally, Engelhard and Kollins (2019) emphasized that individuals with ADHD exhibit heightened sensitivity to screen media technology, which may contribute to intensified symptoms, and they underscored the necessity for further investigation in this domain.

Collectively, these studies reflect a diverse array of findings, with some demonstrating a clear association between increased screen time and the worsening of ADHD symptoms, while others suggest a weaker or negligible correlation. These results underscore the complexity of the interplay between screen time and ADHD, indicating that factors such as the type of screen exposure, age, and other lifestyle variables may significantly influence outcomes. A summary of the key findings from the articles revealing an association between ADHD and screen time is presented in Table 2.

## Discussion

### Pathophysiology of ADHD

The precise etiology of Attention-Deficit/Hyperactivity Disorder (ADHD) remains poorly understood; however, several theories have been proposed to elucidate the disorder. Research indicates that ADHD may be influenced by the functioning of certain neurotransmitters, specifically dopamine and noradrenaline, alongside structural variations in the brain. A notable correlation between diminished dopamine levels and ADHD has been demonstrated through advanced imaging techniques, such as Positron Emission Tomography (PET) scans (Mehta, 2019). Individuals with ADHD frequently exhibit difficulties in impulse control, attributed to abnormalities in these neurochemical pathways. Neuroimaging studies employing Magnetic Resonance Imaging (MRI) have revealed structural differences in specific brain regions, including the prefrontal cortex, cerebellum, and various subcortical structures, in individuals diagnosed with ADHD (Srichawla et al., 2022). Additionally, Diffusion Tensor Imaging (DTI) has illustrated alterations in cortical morphology among adolescents with ADHD. Functional Magnetic Resonance Imaging (fMRI) scans conducted during tasks requiring attention and self-regulation have revealed reduced activation in particular neural pathways among individuals with ADHD (Hart et al., 2013).

### Gender and Age-Related Factors

ADHD affects both males and females; however, prevalence rates indicate that it is more frequently diagnosed in boys. In contrast, girls with ADHD are predominantly diagnosed with the inattentive subtype. While there are few significant differences between boys and girls diagnosed with ADHD, adolescent females tend to struggle more with issues related to self-confidence and adversity compared to their male counterparts (Rucklidge, 2010). Furthermore, girls with ADHD may experience elevated levels of depression and anxiety; however, they typically exhibit less physical aggression and fewer overt behavioral manifestations. Varied factors, such as impulsivity, academic performance, social skills, fine motor skills, parental education, and mood, generally do not exhibit substantial gender differences. Nonetheless, girls with ADHD may face increased challenges in cognitive processing, display reduced hyperactivity, and demonstrate fewer externalizing behaviors than boys with the disorder. It is difficult to ascertain whether these observed differences stem from biases in the referral process for evaluation or treatment (Gaub and Carlson, 1997).

Girls with ADHD may not be as readily identified during the diagnostic process and are often prescribed medication only if they exhibit pronounced external symptoms of the disorder (Mowlem et al., 2019). Among the general population, children frequently display behaviors consistent with ADHD—such as inattention, hyperactivity, and impulsivity—particularly after extensive exposure to screens, including television and video games (Nikkelen et al., 2014). Such exposure can result in the development of addictive behaviors regarding screen time or an inability to function without electronic devices. Notably, boys are more likely to develop an addiction to video games, whereas girls may gravitate more towards social networking platforms (Andreassen et al., 2016). Excessive screen time, particularly among preschoolers, has been linked to exacerbated attention problems (Tamana et al., 2019). Specifically, children who engage with screens for more than two hours daily by the age of five are at an increased risk of developing behavioral issues. The association between screen time and behavioral problems is more pronounced than other factors, including sleep quality, parental stress, and socioeconomic status. It is crucial to educate parents, especially during the preschool years, about the importance of limiting screen time and promoting physical activities to mitigate attention difficulties in children. Some studies have suggested that children who engage in conversations on mobile phones or play games on such devices may be at a heightened risk for ADHD. It remains unclear whether these children utilize their phones more frequently due to pre-existing ADHD symptoms or if excessive phone use contributes to the onset of ADHD (Zalsman and Shilton, 2016).

### Genetic Factors

Attention Deficit Hyperactivity Disorder (ADHD) is a prevalent neurodevelopmental disorder with a profound genetic component, as evidenced by studies indicating that heritability accounts for approximately 74% of its development. Genetic investigations have identified several gene variants associated with ADHD; however, these variants elucidate only a portion of the disorder’s heritability. Researchers have engaged in extensive studies of familial patterns and genetic linkage to pinpoint specific genes linked to ADHD, discovering that each DNA risk variant exerts a modest effect on the disorder. Moreover, genome-wide association studies (GWAS) have successfully identified genetic loci statistically correlated with ADHD. Notably, molecular genetic findings have predominantly emerged from research exploring specific genes and their functional implications (Faraone and Larsson, 2019). Substantial evidence consistently connects ADHD to certain dopamine receptor genes. Additionally, recent literature indicates that specific genetic variations may contribute to conduct problems in individuals diagnosed with ADHD (Thapar and Stergiakouli, 2008).

### Comorbidities of ADHD and Screen Time

A 2023 study provides compelling evidence for the relationship between ADHD symptoms and screen time among children (Waller et al., 2023). This research, which analyzed data from 68,634 children aged 5 to 17 years from the National Survey of Children’s Health, found that approximately 10% of participants were diagnosed with ADHD. Factors such as male gender, healthcare coverage, and increased levels of depression and anxiety were shown to elevate the likelihood of an ADHD diagnosis. The study underscored the association between ADHD and various factors, including screen usage and television viewing, in addition to other environmental influences.

Parents reported exacerbation of their children’s ADHD symptoms corresponding to increased screen time (Pickren et al., 2022). Another investigation identified a tenuous link between the number of hours spent watching television and ADHD symptoms, particularly in urban settings (Tan and Zhou, 2022). Furthermore, excessive engagement with video games (Masi et al., 2021) and social media (Shuai et al., 2021) was shown to amplify ADHD symptoms (Thorell et al., 2022). Notably, children exhibiting ADHD symptoms frequently demonstrate unhealthy lifestyle patterns, such as prolonged sedentary behavior and poor dietary choices (Tong et al., 2016). In contrast, the research found that obesity was not significantly associated with ADHD, contrary to the findings of some prior studies (Lingineni et al., 2012).

The study highlighted a relationship between ADHD, suboptimal academic performance, and poor sleep quality in connection with screen time (Paulich et al., 2021). Furthermore, ADHD is frequently accompanied by several comorbid neurological and psychiatric disorders. More than 60% of individuals with ADHD also exhibit one or more additional disorders, such as oppositional defiant disorder, conduct disorder, anxiety, depression, and tic disorders. Between 20% and 60% of children with ADHD struggle with learning disabilities, with dyslexia and dyscalculia being the most prevalent (Faraone and Larsson, 2019), and dyslexia occurring more frequently in boys (Lingineni et al., 2012). This condition hinders the ability to learn by impacting reading and spelling proficiency. The underlying mechanisms contributing to these associations remain to be fully elucidated (Faraone and Larsson, 2019).

## Conclusion

This comprehensive review suggests a credible association between screen time and ADHD. While direct causation remains to be definitively established, substantial evidence elucidates a significant correlation. The relationship is multifaceted and influenced by various factors. Excessive screen time adversely affects ADHD symptoms by disrupting sleep, hindering concentration, diminishing physical activity, and reducing social interactions. To mitigate these challenges, it is imperative for healthcare providers, parents, and educators to recognize the potential risks associated with excessive screen use. Given the limitations of current data, further extensive research is warranted. This study enhances our understanding of the issue and emphasizes the need for robust guidelines to regulate screen time and effectively manage ADHD.

## Data Availability

All data produced in the present work are contained in the manuscript

## References

1. Andreassen, C. S., et al. (2016). The relationship between addictive use of social media and video games and symptoms of psychiatric disorders: A large-scale cross-sectional study. Psychology of Addictive Behaviors, 30(2), 252.

2. Attygalle, U. R., et al. (2020). Migraine, attention deficit hyperactivity disorder and screen time in children attending a Sri Lankan tertiary care facility: are they associated? BMC Neurology, 20(1), 275. doi:10.1186/s12883-020-01855-5

3. Brem, S., et al. (2014). The neurobiological link between OCD and ADHD. ADHD Attention Deficit and Hyperactivity Disorders, 6, 175–202.

4. Cavalli, E., et al. (2021). Screen exposure exacerbates ADHD symptoms indirectly through increased sleep disturbance. Sleep Medicine, 83, 241–247.

5. Chassiakos, Y. L., et al. (2016). Children and adolescents and digital media. Pediatrics, 138(5).

6. Drechsler, R., et al. (2020). ADHD: Current concepts and treatments in children and adolescents. Neuropediatrics, 51(05), 315–335.

7. Engelhard, M. M., & Kollins, S. H. (2019). The many channels of screen media technology in ADHD: A paradigm for quantifying distinct risks and potential benefits. Current Psychiatry Reports, 21, 1–10.

8. Faraone, S. V., & Larsson, H. (2019). Genetics of attention deficit hyperactivity disorder. Molecular Psychiatry, 24(4), 562–575.

9. Gaub, M., & Carlson, C. L. (1997). Behavioral characteristics of DSM-IV ADHD subtypes in a school-based population. Journal of Abnormal Child Psychology, 25, 103–111.

10. Hansen, J., et al. (2022). Physical activity, screen time, and sleep: Do German children and adolescents meet the movement guidelines? European Journal of Pediatrics, 181(5), 1985–1995.

11. Hart, H., et al. (2013). Meta-analysis of functional magnetic resonance imaging studies of inhibition and attention in attention-deficit/hyperactivity disorder: exploring task-specific, stimulant medication, and age effects. JAMA Psychiatry, 70(2), 185-198. doi:10.1001/jamapsychiatry.2013.277

12. Hill, M. M., et al. (2020). Screen time in 36-month-olds at increased likelihood for ASD and ADHD. Infant Behavior and Development, 61, 101484. doi:10.1016/j.infbeh.2020.101484

13. Klassen, A. F., et al. (2004). Health-related quality of life in children and adolescents who have a diagnosis of attention-deficit/hyperactivity disorder. Pediatrics, 114(5), e541–e547.

14. Levelink, B., et al. (2021). The Longitudinal Relationship Between Screen Time, Sleep and a Diagnosis of Attention-Deficit/Hyperactivity Disorder in Childhood. Journal of Attention Disorders, 25(14), 2003–2013. doi:10.1177/1087054720953897

15. Lingineni, R. K., et al. (2012). Factors associated with attention deficit/hyperactivity disorder among US children: Results from a national survey. BMC Pediatrics, 12, 1–10.

16. Lissak, G. (2018). Adverse physiological and psychological effects of screen time on children and adolescents: Literature review and case study. Environmental Research, 164, 149–157.

17. Masi, L., et al. (2021). Video games in ADHD and non-ADHD children: Modalities of use and association with ADHD symptoms. Frontiers in Pediatrics, 9, 632272.

18. Mehta, T. R., et al. (2019). Neurobiology of ADHD: A review. Current Developmental Disorders Reports, 6, 235–240.

19. Montagni, I., et al. (2016). Association of screen time with self-perceived attention problems and hyperactivity levels in French students: a cross-sectional study. BMJ Open, 6(2), e009089. doi:10.1136/bmjopen-2015-009089

20. Mowlem, F. D., et al. (2019). Sex differences in predicting ADHD clinical diagnosis and pharmacological treatment. European Child & Adolescent Psychiatry, 28(4), 481–489. doi:10.1007/s00787-018-1211-3

21. Nikkelen, S. W., et al. (2014). Media use and ADHD-related behaviors in children and adolescents: A meta-analysis. Developmental Psychology, 50(9), 2228.

22. Paulich, K. N., et al. (2021). Screen time and early adolescent mental health, academic, and social outcomes in 9- and 10- year old children: Utilizing the Adolescent Brain Cognitive Development (ABCD) Study. PLoS One, 16(9), e0256591. doi:10.1371/journal.pone.0256591

23. Peppers, K. H., et al. (2016). An intervention to promote sleep and reduce ADHD symptoms. Journal of Pediatric Health Care, 30(6), e43–e48.

24. Pickren, S. E., et al. (2022). Impact of COVID-19 on Children’s Attention Deficit Hyperactivity Disorder Symptomology, Daily Life, and Problem Behavior During Virtual Learning. Mind, Brain, and Education, 16(4), 277–292. doi:10.1111/mbe.12337

25. Rucklidge, J. J. (2010). Gender differences in attention-deficit/hyperactivity disorder. Psychiatric Clinics, 33(2), 357–373.

26. Shuai, L., et al. (2021). Influences of digital media use on children and adolescents with ADHD during COVID-19 pandemic. Globalization and Health, 17, 1–9.

27. Soares, P. S., et al. (2022). Is screen time throughout adolescence related to ADHD? Findings from 1993 Pelotas (Brazil) birth cohort study. Journal of Attention Disorders, 26(3), 331-339.

28. Srichawla, B. S., et al. (2022). Attention Deficit Hyperactivity Disorder and Substance Use Disorder: A Narrative Review. Cureus, 14(4), e24068. doi:10.7759/cureus.24068

29. Tamana, S. K., et al. (2019). Screen-time is associated with inattention problems in preschoolers: Results from the CHILD birth cohort study. PLoS One, 14(4), e0213995.

30. Tan, T. X., & Zhou, Y. (2022). Screen Time and ADHD Behaviors in Chinese Children: Findings From Longitudinal and Cross-Sectional Data. Journal of Attention Disorders, 26(13),

31. Thapar, A., & Stergiakouli, E. (2008). An Overview on the Genetics of ADHD. Xin li xue bao. Acta Psychologica Sinica, 40(10), 1088.

32. Thoma, V. K., et al. (2020). Media use, sleep quality, and ADHD symptoms in a community sample and a sample of ADHD patients aged 8 to 18 years. Journal of Attention Disorders, 24(4), 576–589.

33. Thorell, L. B., et al. (2022). Longitudinal associations between digital media use and ADHD symptoms in children and adolescents: A systematic literature review. European Child & Adolescent Psychiatry. Advance online publication. doi:10.1007/s00787-022-02120-7

34. Tong, L., et al. (2016). Attention-Deficit/Hyperactivity Disorder and Lifestyle-Related Behaviors in Children. PLoS One, 11(9), e0163434. doi:10.1371/journal.pone.0163434

35. Vaidyanathan, S., et al. (2021). Screen time exposure in preschool children with ADHD: A cross-sectional exploratory study from South India. Indian Journal of Psychological Medicine, 43(2), 125–129.

36. Waller, F., et al. (2023). Screen use: Its association with caregiver mental health, parenting, and children’s ADHD symptoms. Family Relations.

37. Yang, A., et al. (2022). Longer screen time utilization is associated with the polygenic risk for Attention-deficit/hyperactivity disorder with mediation by brain white matter microstructure. EBioMedicine, 80, 104039. doi:10.1016/j.ebiom.2022.104039

38. Zalsman, G., & Shilton, T. (2016). Adult ADHD: A new disease? International Journal of Psychiatry in Clinical Practice, 20(2), 70–76.

